# LLM-enabled Natural History Study Analysis to Support Rare Disease Research

**DOI:** 10.64898/2026.09.17.26363303

**Authors:** Kevin Li, Eric Sid, Qian Zhu

**Author notes:** **Corresponding author: Qian Zhu,**.

## Abstract

**Background:** Rare diseases affect an estimated 300 million people worldwide, yet the research needed to guide diagnosis and treatment is often fragmented across multiple unstructured literature sources. Natural history studies (NHS) are a key source of this evidence, but manually extracting structured information from NHS publications can be tedious and does not scale.

**Methods:** We have developed a proof-of-concept for an information extraction pipeline testing three open-source large-language models (LLMs) – Athena-v3-AWQ, Google’s Gemma3-27B, and Meta’s Llama-3.1-70B-Instruct, to extract key NHS characteristics from PubMed abstracts curated from a Chan Zuckerberg Initiative disease research state model corpus (302 gold-standard and 8,338 full-corpus abstracts), and compared the models on efficiency, extraction completeness, and expert-rated accuracy.

**Results:** All three models processed abstracts with success rates exceeding 99%. However, Gemma achieved the best overall performance, with the highest expert-rated accuracy (68.0% of outputs rated “good” vs. 36.0% for Llama and 10.0% for Athena) and the fastest runtime on the full corpus (∼16 minutes for 3,547 abstracts), despite Llama scoring higher on the automated Token F1 metric (0.874 vs. 0.723), highlighting a divergence between automated and human evaluation. Athena’s lower performance was largely attributable to verbatim copying rather than synthesis of extracted content.

**Conclusions:** These findings illustrate how locally deployed open-source LLMs can extract structured NHS characteristics at scale, thus supporting their use to accelerate evidence synthesis in rare disease research.

## Background

A rare disease is defined as a condition that affects fewer than 200,000 people in the U.S.[1] However, there are more than 10,000 classified conditions[2] that affect an estimated 300 million people worldwide[3], including roughly 30 million Americans.[4] Despite such a large population, there are many challenges in researching and treating these diseases and the patients who are affected. Approximately 95% of rare diseases have no approved treatment[5], and patients often undergo multiple clinical visits before receiving an accurate diagnosis. This delay is made more difficult to deal with due to the fragmented nature of rare disease data. Patient cohorts can be small and geographically dispersed, making it difficult to identify disease mechanisms and treatment responses.[6] Rare diseases also disproportionally affect children[3], which can result in life-threatening or fatal symptoms, necessitating an urgent improvement in research infrastructure. Such challenges emphasize a need for evidence-based approaches to understand disease biology and accelerate the development of new therapies.

Natural history studies (NHS) address this information gap by identifying demographic, genetic, environmental, and clinical variables that characterize a patient population over time.[7] While large randomized clinical trials are not feasible for rare diseases with limited populations, NHS serves as a trusted alternative for understanding how a disease manifests and responds to treatment.[8] Regulatory agencies like the U.S. Food and Drug Administration and the European Medicines Agency recognize real-world evidence-based analyses as capable of establishing natural endpoints against which therapeutic efficacy can be measured.[7,9] NHS also plays a role in identifying patient subgroups and informing clinical trial design, adding to the versatility of these studies in mapping rare disease progression.[10] However, the insights embedded in NHS can be spread across a multitude of different resources, including publications and clinical trials, and the complexity of their reporting formats can make it difficult to extract and analyze key findings from unstructured text at large.

Recent advances in large language models (LLMs) enable automated information extraction, making it easier to aggregate key study characteristics and analyze disease features at scale. Early approaches have relied on rule-based methods that require extensive curation and can struggle to adapt to texts of differing topics.[11] Machine learning (ML) techniques are becoming increasingly prevalent in data analysis, with methods such as support vector machines and conditional random fields enabling more flexible applications of natural language processing and relation extraction from biomedical text.[12,13] However, they still depend on manually created features and annotated training data, limiting their ability to generalize across the complex language found in clinical literature. Building on these foundations, deep learning (DL) architectures like BioBERT and MSR BiomedBERT further improve extraction performance by leveraging pre-trained models trained on medical texts to domain-specific publications, achieving high performance on benchmark tasks like named entity recognition and question answering.[14,15] In general, large language models (LLMs) have developed powerful capabilities to obtain structured outputs from unstructured text without program-dependent fine-tuning.[16] These capabilities make LLMs suitable for biomedical extraction tasks where annotated training data is scarce, as is often the case in rare disease research.[17]

In this study, we aimed to establish a proof-of-concept for applying LLMs to extract information from a curated set of NHS-based PubMed abstracts to identify essential elements that may correlate with the development and outcomes of a rare disease. In addition, we compared efficiency and accuracy among several open-source LLMs for this research use case. Through our research, we found that LLMs are a suitable tool for analyzing NHS data to support rare disease research.

## Methods

### 1. NHS corpus description

We obtained a corpus of PubMed abstracts from the Chan Zuckerberg Initiative Disease Research State Model (CZI DRSM) corpus.[18] The corpus included two sets of abstracts: one with “Labeling_State” set to “gold standard,” curated by the Chan Zuckerberg curation team, and another with “Labeling_State” set to “Labeled,” curated by the Centaur Labs curation service.[18] For each abstract, it was further grouped into one of four “Label Categories” shown in Table 1, based on their relevance to natural history studies (NHS) in rare disease research.[18]

**Table 1.** Four categories applied to label each PubMed abstract in the CZI DRSM corpus.

| Label Categories |
| --- |
| '-1 - the paper is not a primary experimental study in rare disease' |
| '0 - the study is not directly investigating the natural history of a disease' |
| '1 - the study includes some elements a natural history but not as its primary contribution' |
| '2 - the study's primary contribution centers on observing the time course of a rare disease' |

To ensure NHS-related information was presented in the abstracts, in this preliminary study, we focused only on the abstracts labeled “2 - the study’s primary contribution centers on observing the time course of a rare disease,” shown in Table 1. For the initial model testing and prompt engineering step, we used the “gold standard” corpus. After optimizing our extraction framework, we expanded our analysis to the entire corpus labeled “2”.

### 2. LLM models used

Three open-source LLMs, Athena-v3-AWQ, Google’s Gemma3-27b, and Meta’s Llama-3.1-70B-Instruct, were available within the NCATS computational infrastructure to develop and evaluate our information extraction pipeline. In the remainder of this manuscript, Athena-v3-AWQ, Google Gemma3-27B, and Meta Llama-3.1-70B-Instruct are referred to as Athena, Gemma, and Llama, respectively.

- Athena is a community-developed 13-billion-parameter model that employs Activation-aware Weight Quantization (AWQ) at 4-bit precision, identifying which 1% of a model’s weights are most critical for accurate outputs and scaling them to protect them from quantization rounding errors, enabling accurate use after 4-bit compression.[19,20] This enables faster inference in lower-memory environments.[20]
- Gemma is a 27-billion-parameter model released by Google in March 2025 and has been touted for large-scale, high-context tasks at efficient speeds.[21]
- Llama is a 70-billion-parameter model released by Meta in July 2024, designed for complex reasoning tasks at a level comparable to closed-source models.[22]

The three models vary in architecture and design intent, providing a range of performance profiles that enable meaningful comparisons of extraction quality, completeness, and runtime efficiency across models with differing capabilities. All three were available within the existing NCATS computational infrastructure, making them practical choices for deployment without additional resource procurement. These open-source models represent viable alternatives to proprietary systems for institutions seeking cost-effective, locally deployable solutions for biomedical text analysis. This diversity in model characteristics allows our evaluation to produce findings that are broadly informative for researchers selecting LLMs for similar biomedical information extraction tasks.

In this study, we demonstrated a proof-of-concept for using an LLM to identify critical elements in NHS. Working with domain experts, we first defined a set of NHS-focused research questions to identify key information reported in NHS. We then applied and compared multiple open-source LLMs to analyze NHS abstracts and evaluate their ability to accurately answer those predefined questions. The study workflow is shown in Figure 1.

**Figure 1.**
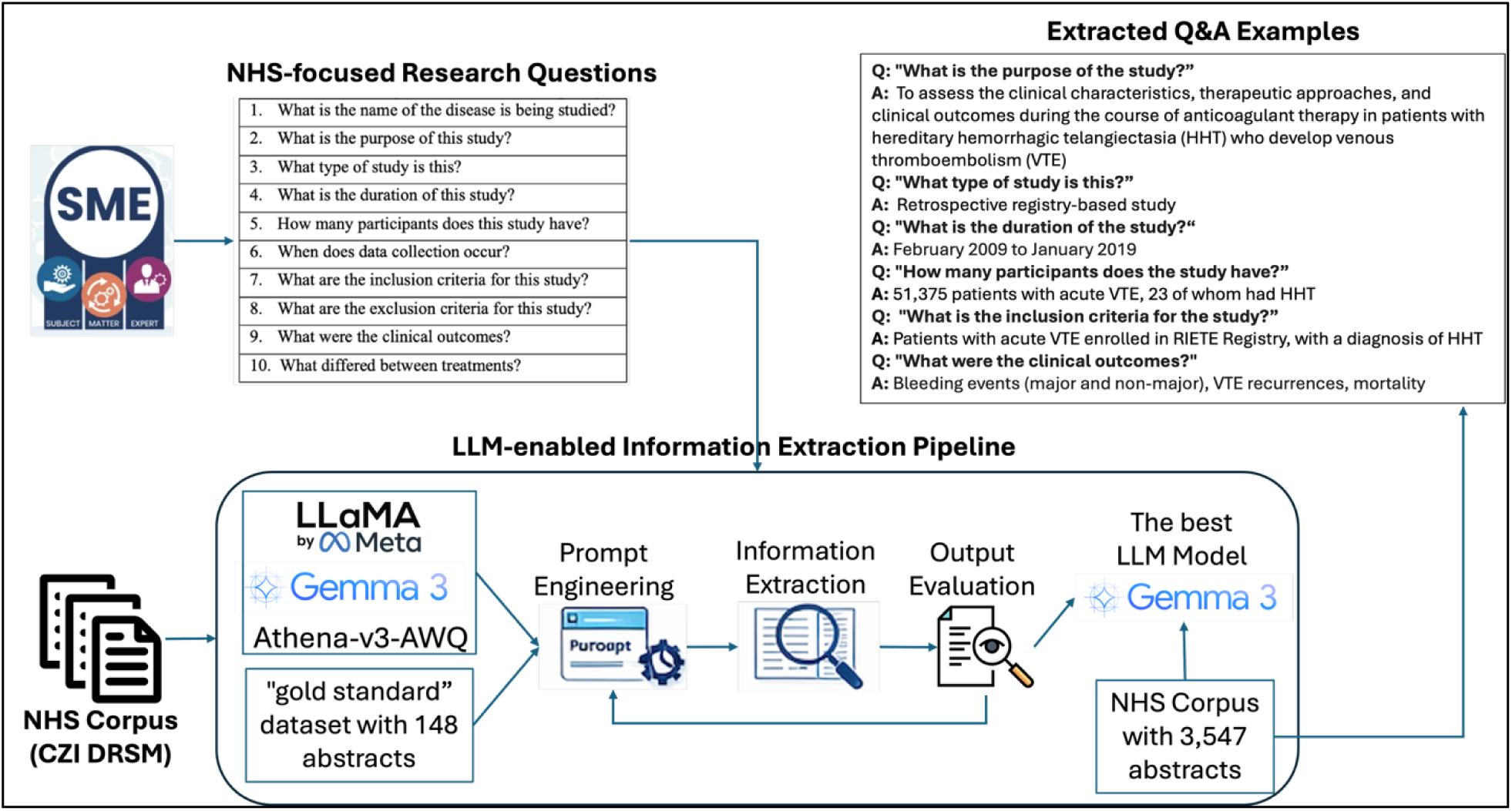
Study workflow for LLM enabled NHS analysis.

### 3. NHS-focused research question definition

We worked with our rare disease expert (co-author, Eric Sid, MD) from the Division of Rare Diseases Research Innovations (DRDRI) at the National Center for Advancing Translational Sciences (NCATS)/National Institutes of Health (NIH) to identify the critical elements rare disease researchers are mostly interested in from NHS. Through the discussion and his guidance, we formulated 10 scientific questions, listed as “NHS-focused research questions” in Figure 1, which reflect the specific information they sought from the NHS. To facilitate implementation by the LLM in the subsequent step, we translated these 10 questions into 11 precisely defined characteristics shown in Table 2.

**Table 2.**
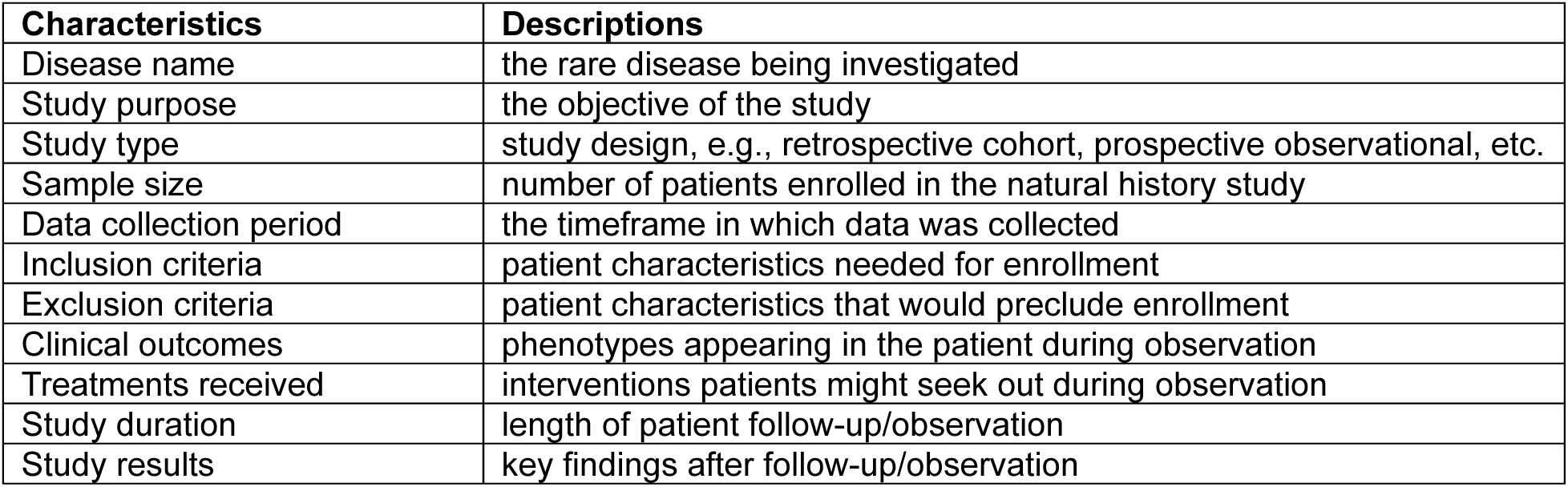
11 scientific characteristics translated from NHS-focused research questions.

| Characteristics | Descriptions |
| --- | --- |
| Disease name | the rare disease being investigated |
| Study purpose | the objective of the study |
| Study type | study design, e.g., retrospective cohort, prospective observational, etc. |
| Sample size | number of patients enrolled in the natural history study |
| Data collection period | the timeframe in which data was collected |
| Inclusion criteria | patient characteristics needed for enrollment |
| Exclusion criteria | patient characteristics that would preclude enrollment |
| Clinical outcomes | phenotypes appearing in the patient during observation |
| Treatments received | interventions patients might seek out during observation |
| Study duration | length of patient follow-up/observation |
| Study results | key findings after follow-up/observation |

### 4. Prompt engineering

In this study, we aimed to develop an information extraction pipeline using LLMs. Prompt engineering was performed iteratively to progressively improve the accuracy and consistency of LLM outputs across all 11 selected study characteristics shown in Table 2. The process consists of multiple stages informed by manual inspection of model outputs against the source abstracts. In the initial stage, a base prompt (as shown in Figure 2) was constructed to list the target study characteristics and instruct the model to return responses as a JSON object, using “N/A” for characteristics not reported in the abstract.

**Figure 2.**
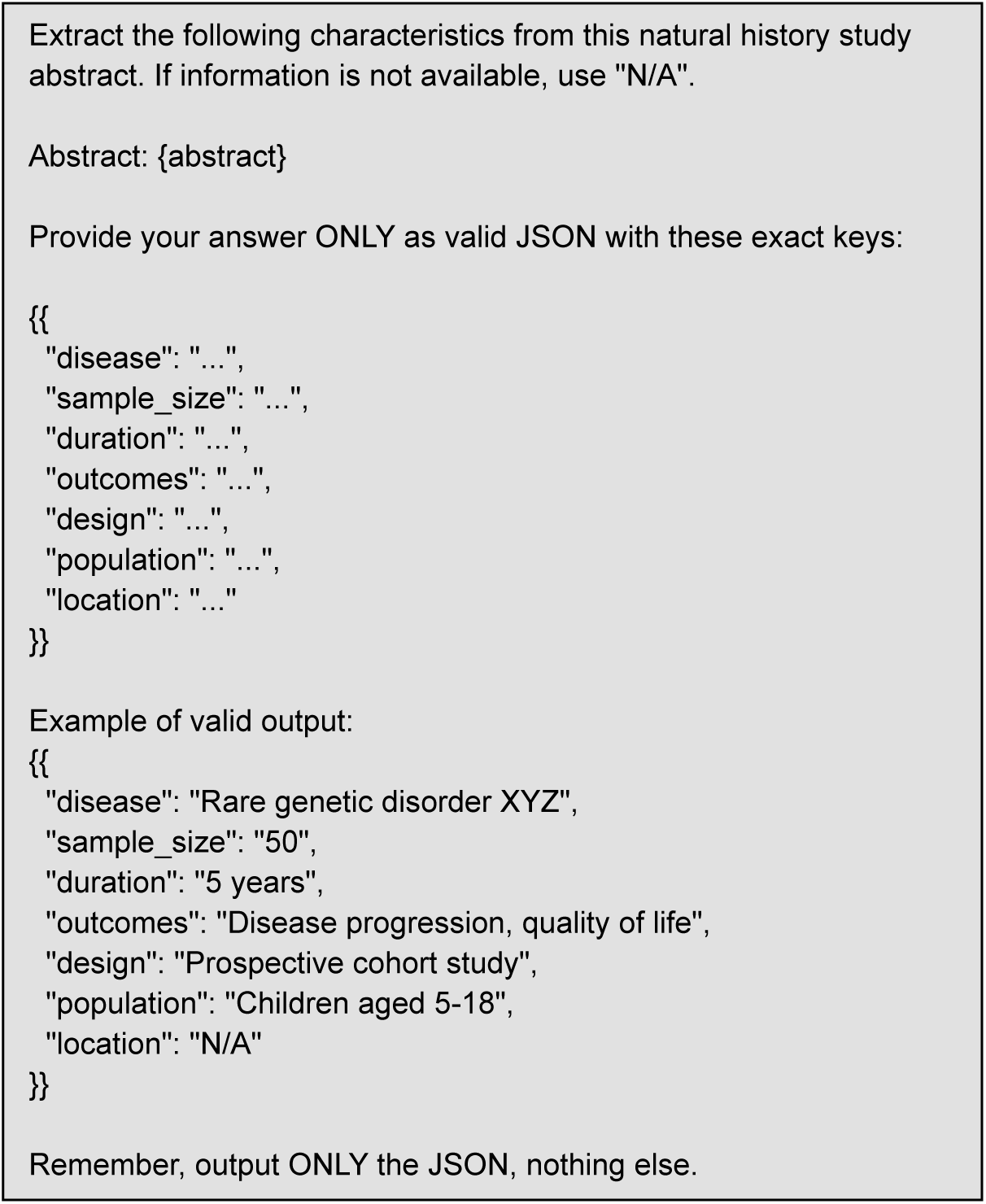
First prompt used for data extraction on natural history abstracts.

Although models generally followed the requested output structure, outputs showed inconsistent interpretation of the characteristics and frequent misclassification of extracted content. For example, when applied to an abstract describing a pediatric primary sclerosing cholangitis (PSC) cohort,[23] the initial prompt returned “Liver transplantation, cirrhosis, biochemical abnormalities, fibrosis” for the undefined “outcomes” field, conflating a treatment (liver transplantation) with clinical outcomes while omitting inportant disease-specific findings reported in the abstract, such as autoimmune sclerosing cholangitis, METAVIR fibrosis staging, and inflammatory bowel disease. In addition, the initial prompt schema lacked dedicated fields for study purpose, inclusion and exclusion criteria, and results, limiting the comprehensiveness and graularity of the extracted information. To address these issues, we refined the prompt by adding an explicit definition for each characteristic, thereby constraining model interpretation and reducing ambiguity. Example outputs were also incorporated to guide consistent phrasing, particularly for fields requiring standardized or stable formats for downstream analysis.

Additional refinements were introduced iteratively based on the error patterns observed during prompt evaluation. For instance, the definition of “clinical outcomes” was revised to explicitly exclude treatment related information, such as treatment efficacy, and instead focus on observed disease phenotypes, clinical manifestations, symptoms and other outcomes directly associated with the rare disease. The definition of “Treatments received” was broadened to include all interventions received by patients, rather than only the treatment compared across groups. For instance, when applied to the same above mentioned PSC abstract,[23] the *clinical_outcomes* field evolved from a narrative-style summary that combined disease features with statistical detail to a concise, comma-separated list of phenotype phrases consistent with the refined definition (e.g., “autoimmune sclerosing cholangitis (ASC), exclusive small duct PSC, F3-F4 METAVIR stage, inflammatory bowel disease (IBD)”). Similarly, the *treatments_received* field replaced the earlier “differences between treatments” formulation, which conflated treatment information with comparative statistical findings (e.g., “Liver transplantation rate was lower in patients with PSC and IBD than in those without IBD (2% vs 18%, P=0.01)”), with a concise and standardized statement of the intervention received (e.g., “liver transplant”). Additional instructions were incorporated to improve JSON validity and minimize formatting errors that could interfere with automated parsing of the extracted results. For fields containing multiple entries, the prompt specified a comma-separated formate and included representative examples to clarify the expected level of granularity and specificity. The final optimized prompt shown in Figure 3, was then applied consistently across all three LLMs to ensure a fair and standarized comparison of model performance.

**Figure 3.**
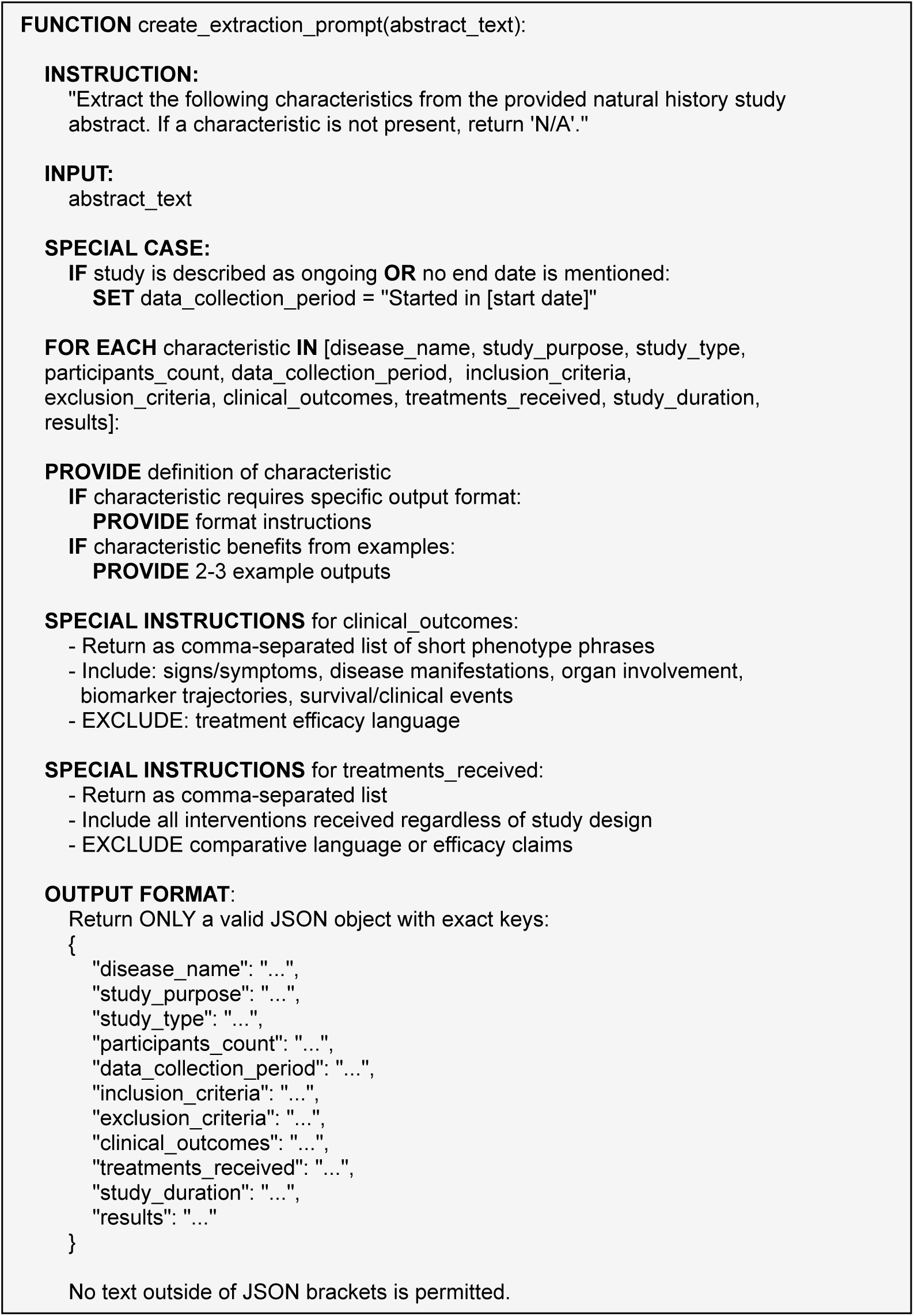
Format of the final prompt used for information extraction.

### 5. Information extraction

#### 5.1. LLM model configuration

All three models were deployed with identical configurations to ensure fair comparison. Each model was run across four graphic processing units (GPUs) with 90% GPU memory utilization and a maximum context length of 3,072 tokens. For text generation, a low sampling temperature of 0.1 was applied to promote consistent and deterministic outputs. The maximum output length was set to 2,048 tokens, with a nucleus sampling probability of 0.95. Predefined stop sequences were also specified to ensure that model responses terminated cleanly and could be reliably parsed into valid JSON files. All models were deployed using the vLLM Python library,[24] which supports large-scale inference across multiple GPUs.

#### 5.2. LLM output standardization

To standardize LLM output by aligning with controlled vocabularies, we mapped extracted rare disease names to GARD (NCATS Genetic and Rare Diseases Information Center),[5] “Clinical outcomes” to Human Phenotype Ontology (HPO) via its API,[25] and “Treatments received” to RxNorm via its API,[26] respectively.

#### 5.3. LLM model performance evaluation

To assess model performance, each LLM was evaluated using 11 consecutive runs on the same set of abstracts. The first run was used to measure model loading time and initial inference runtime, while the subsequent 10 runs were averaged to estimate typical runtime performance.

Model performance was evaluated using the following criteria:

- Extraction completeness: the number and percentage of abstracts for which the model successfully generated complete structured outputs.
- Output quality: qualitative assessment of the extracted information by comparing the model-generated outputs with the corresponding abstract text.

To further assess extraction accuracy, we randomly selected 50 abstracts along with their corresponding outputs generated by each of the three LLMs. These outputs were independently reviewed by our rare disease expert (Eric Sid, MD), who qualitatively evaluated the accuracy, completeness, and clinical relevance of the extracted information.

## Results

## 1. Results on the NHS Corpus

Table 3 and Table 4 summarize the distribution of abstracts by NHS-relevance label category within the gold-standard dataset (302 PMIDs, or PubMed identifier) and the full CZI DRSM corpus (8,338 PMIDs), respectively. An example of LLM-generated extraction output is shown in Figure 4.

**Figure 4.**
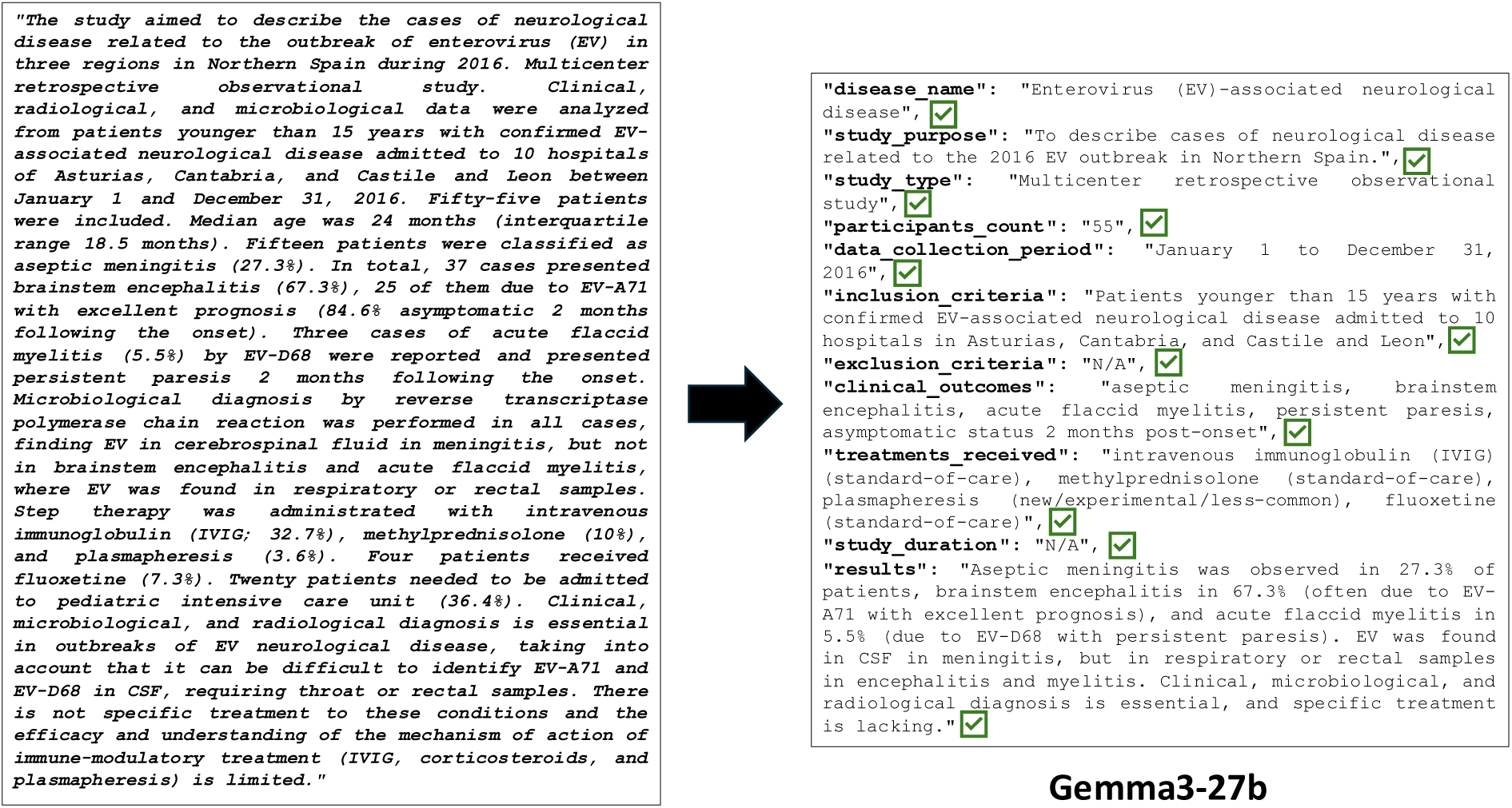
Example of LLM extraction output on an example of NHS based abstract.

**Table 3.** Four categories of PubMed articles from the “Gold Standard” set.

| Label Categories | Count of PMID |
| --- | --- |
| '-1 - the paper is not a primary experimental study in rare disease' | 12 |
| '0 - the study is not directly investigating the natural history of a disease' | 112 |
| '1 - the study includes some elements a natural history but not as its primary contribution' | 30 |
| '2 - the study's primary contribution centers on observing the time course of a rare disease' | 148 |
| Total of PMID | 302 |

**Table 4.** Four categories of PubMed articles from the CZI DRSM corpus.

| Label Categories | Count of PMID |
| --- | --- |
| '-1 - the paper is not a primary experimental study in rare disease' | 1,988 |
| '0 - the study is not directly investigating the natural history of a disease' | 1,378 |
| '1 - the study includes some elements a natural history but not as its primary contribution' | 1,425 |
| '2 - the study's primary contribution centers on observing the time course of a rare disease' | 3,547 |
| Total of PMID | 8,338 |

## 2. Results on LLM model performance assessment

Across the three evaluated performance metrics: efficiency, completeness, and accuracy, Gemma demonstrated the strongest overall performance for information extraction. It achieved the highest expert-rated accuracy, with 68% of responses rated as good, while also maintaining strong extraction completeness. Although Llama achieved a higher automated Token F1 score (0.874 vs. 0.723 for Gemma), expert evaluation indicated that Gemma provided more accurate and clinically meaningful extracted information.

### 2.1. Efficiency assessment

Runtime performance across all three models is summarized in Table 5. For the 148-gold standard abstract set, Athena demonstrated the shortest average runtime at 1 minute 51.9 seconds, followed by Gemma at 3 minutes 22.6 seconds and Llama at 3 minutes 49.6 seconds. The low standard deviations observed across all models indicate consistent and reproducible runtime performance on this evaluation set. When applied to the full corpus of 3,547 abstracts, the relative runtime performance changed. Gemma achieved the fastest average runtime at 16 minutes 1.6 seconds, followed by Athena at 18 minutes 15.1 seconds. In contrast, Llama required substantially longer processing time, with an average runtime of 40 minutes, 14.4 seconds. These results suggest that while Athena provided the highest efficiency on the smaller gold-standard dataset, Gemma showed improved scalability and processing efficiency when applied to a larger corpus of abstracts.

**Table 5.** Run times and success rates across 148 gold standard abstracts.

| Characteristics | Athena | Gemma | Llama |
| --- | --- | --- | --- |
| **Run time for 148 abstracts on startup** | 2 minutes, 23.4 seconds | 4 minutes, 16.7 seconds | 5 minutes, 12.3 seconds |
| **Average run time for 148 abstracts over 10 extra runs** | 1 minute, 51.9 seconds | 3 minutes, 22.6 seconds | 3 minutes, 49.6 seconds |
| **Success rate** | 1 fail out of 148 | 0 fails out of 148 | 0 fails out of 148 |

### 2.2. Completeness assessment

Completeness was assessed by examining the missing output rate for each extracted characteristic across the 148 gold standard abstracts, as summarized in Table 6. Gemma and Llama successfully generated outputs for all 148 abstracts, whereas Athena failed to extract information from one abstract. A similar pattern was observed in the full corpus analysis (Table 7), where Athena failed on 39 of 3,547 abstracts, while Gemma and Llama completed extraction for all abstracts.

**Table 6.**
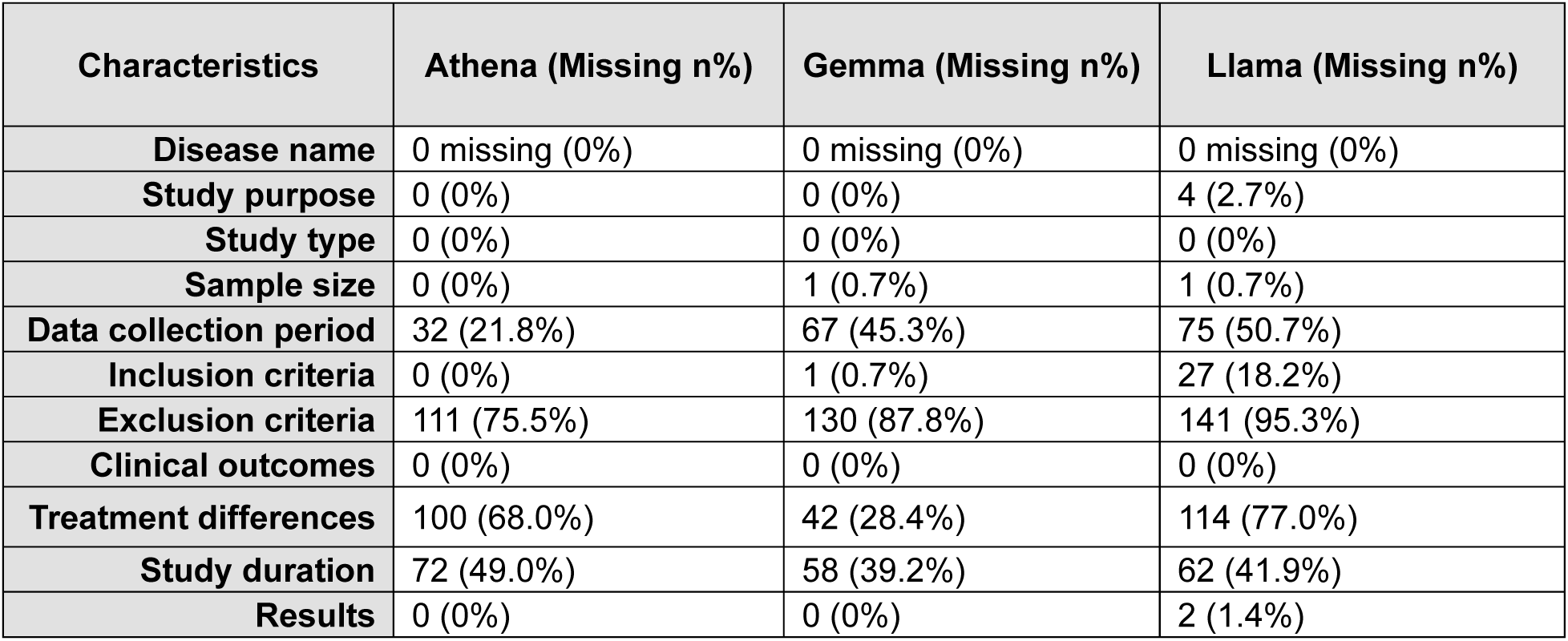
Missing output rates across 148 gold standard abstracts.

| Characteristics | Athena (Missing n%) | Gemma (Missing n%) | Llama (Missing n%) |
| --- | --- | --- | --- |
| **Disease name** | 0 missing (0%) | 0 missing (0%) | 0 missing (0%) |
| **Study purpose** | 0 (0%) | 0 (0%) | 4 (2.7%) |
| **Study type** | 0 (0%) | 0 (0%) | 0 (0%) |
| **Sample size** | 0 (0%) | 1 (0.7%) | 1 (0.7%) |
| **Data collection period** | 32 (21.8%) | 67 (45.3%) | 75 (50.7%) |
| **Inclusion criteria** | 0 (0%) | 1 (0.7%) | 27 (18.2%) |
| **Exclusion criteria** | 111 (75.5%) | 130 (87.8%) | 141 (95.3%) |
| **Clinical outcomes** | 0 (0%) | 0 (0%) | 0 (0%) |
| **Treatment differences** | 100 (68.0%) | 42 (28.4%) | 114 (77.0%) |
| **Study duration** | 72 (49.0%) | 58 (39.2%) | 62 (41.9%) |
| **Results** | 0 (0%) | 0 (0%) | 2 (1.4%) |

**Table 7.** Missing output rates across 3,547 abstracts.

| Characteristic | Athena (Missing n%) | Gemma (Missing n%) | Llama (Missing n%) |
| --- | --- | --- | --- |
| **Disease name** | 0 (0%) | 3 (0.1%) | 19 (0.5%) |
| **Study purpose** | 0 (0%) | 1 (0.0%) | 103 (2.9%) |
| **Study type** | 0 (0%) | 12 (0.3%) | 28 (0.8%) |
| **Sample size** | 30 (0.9%) | 142 (4.0%) | 184 (5.2%) |
| **Data collection period** | 182 (5.2%) | 1,722 (48.5%) | 1,944 (54.8%) |
| **Inclusion criteria** | 92 (2.6%) | 76 (2.1%) | 1,088 (30.7%) |
| **Exclusion criteria** | 1,414 (40.3%) | 3,168 (89.3%) | 3,378 (95.2%) |
| **Clinical outcomes** | 1 (0.0%) | 14 (0.4%) | 105 (3.0%) |
| **Treatments received** | 125 (3.6%) | 1,636 (46.1%) | 1,776 (50.1%) |
| **Study duration** | 868 (24.7%) | 1,212 (34.2%) | 1,539 (43.4%) |
| **Results** | 3 (0.1%) | 0 (0%) | 28 (0.8%) |
| **Total extraction failures** | **39 (1.1%)** | **0 (0%)** | **0 (0%)** |

Across all models, several characteristics demonstrated consistently high extraction completeness, including disease name, study purpose, study type, and clinical outcomes. In contrast, exclusion criteria and data collection period showed the highest missing output rates in both the gold standard and full corpus datasets. These findings suggest that LLMs are generally reliable for extracting core study descriptors but have reduced completeness for characteristics that are less consistently reported or more challenging to identify within PubMed abstracts.

Beyond raw extraction completeness, we assessed the ability of extracted terms to be mapped to standardized controlled vocabulary, as shown in Table 8. GARD and RxNorm mapping rates were comparable across all models, ranging from 70.1%–74.2% and 62.4%–77.6%, respectively. In contrast, HPO mapping rates were substantially lower (28.0-38.2%), likely reflecting the greater specificity and granularity of the HPO vocabulary compared with the broader, free-text phenotype descriptions extracted from abstracts.

**Table 8.**
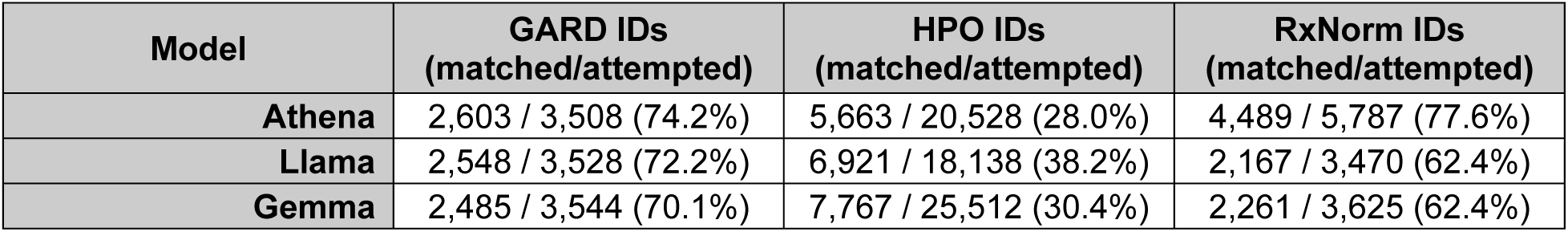
Results on data standardization.

### 2.3. Accuracy assessment

Accuracy was assessed through expert review of model-generated outputs from a randomly selected subset of 50 PubMed abstracts. For each abstract, outputs from all three models were independently evaluated by our rare disease expert (Eric Sid, MD) using a three-point scoring system: 1 = good, 2 = limited, 3 = poor/errors.

Gemma achieved the highest expert-rated performance, with a mean score of 1.40 and 68.0% of responses rated as good. Llama ranked second, with a mean score of 1.76 and 36.0% of responses rated as good. Athena showed substantially lower performance, with a mean score of 2.28 and only 10.0% of responses rated as good. For Gemma and Llama, most remaining errors were associated with fields that had inherently ambiguous boundaries within abstract text, such as study duration and data collection period. These findings suggest that some extraction challenges may be attributable to limitations in how information is reported in abstracts, rather than solely reflecting deficiencies in model performance.

A major factor contributing to Athena’s lower accuracy was its tendency to reproduce text directly from abstracts rather than synthesize the relevant information into concise and structured outputs. For example, in the study results field for an abstract on checkpoint inhibitor (CPI)-related toxicity in older patients,[27] Athena generated an output that closely resembled the original text from the abstract, while Gemma and Llama generated concise summaries capturing the key findings. Gemma summarized the results as: “*CPI use in older patients was not associated with more high-grade toxicity. A positive G8 screening predicted hospital admissions (P = 0.031) and risk of death (P = 0.01). Implementation of G8 screening should be considered in the context of CPI.*” Llama similarly summarized the finding as: *”the use of CPI in older patients was not associated with more high-grade toxicity, a positive G8 screening predicted hospital admissions and risk of death*”.

This pattern was also observed across other extracted characteristics. For example, in a study investigating the distribution of lesions across laryngeal anatomic regions in patients with recurrent respiratory papillomatosis (RRP),[28] Athena again returned text closely mirroring the original abstract for the clinical outcomes field: “*Papilloma locations were recorded using a 21-region laryngeal schematic. Multivariate analyses by anatomic subsite were conducted for the entire population and for subgroups stratified by sex, age, and proton pump inhibitor (PPI) usage. Heat maps were generated, hierarchically color coding the anatomic distribution of disease.*” By comparison, Gemma produced a more concise summary, “*Papilloma locations, number of affected sites, disease prevalence in laryngeal subsites*”, while Llama summarized the outcome as: “*Distribution of RRP lesions across laryngeal anatomic regions*”. These examples illustrate that Athena frequently preserved the original wording and narrative structure of abstracts rather than extracting the intended concepts and converting them into concise, standardized representations.

As a supplementary measure of extraction consistency, automated F1 scores were calculated using a majority-vote framework, as summarized in Table 9. Llama achieved the highest performance, with a Token F1 score of 0.874 and an exact-match F1 score of 0.838, followed by Gemma with a Token F1 score of 0.723 and an exact-match F1 score of 0.695. Athena showed the lowest automated performance, with a Token F1 score of 0.545 and an exact-match F1 score of 0.514. These statistical results were generally consistent with expert assessment, particularly in identifying Athena as the lowest performing model overall.

**Table 9.** Comparative evaluation of three LLMs based on automated metrics and expert review.

| Metric | Athena | Gemma | Llama |
| --- | --- | --- | --- |
| **Human evaluation (50 abstracts manually reviewed)** |  |  |  |
| Mean score | 2.28 | 1.40 | 1.76 |
| Good responses (%) | 10.0% | 68.0% | 36.0% |
| Limited responses (%) | 52.0% | 24.0% | 52.0% |
| Poor responses (%) | 38.0% | 8.0% | 12.0% |
| **Automated Metrics (Majority vote reference, same 50-abstract subset)** |  |  |  |
| Exact match F1 (structured fields) | 0.514 | 0.695 | 0.838 |
| Token F1 (free-text fields) | 0.545 | 0.723 | 0.874 |
| BERTScore (free-text fields) | 0.958 | 0.977 | 0.991 |

## Discussion

We established a proof-of-concept workflow for automated information extraction from natural history study related abstracts using open-source large language models. All three models (Athena, Gemma, and Llama) were able to process abstracts and extract information with very high success rates exceeding 99%. However, our findings also reveal meaningful differences among the models in runtime efficiency, extraction completeness, and output quality. These differences should be carefully considered when selecting and deploying LLM-based approaches for biomedical literature extraction.

Gemma emerged as the most suitable model for large-scale deployment, demonstrating the best overall balance among runtime efficiency, extraction completeness, and output quality. It processed the full corpus of 3,547 abstracts in approximately 16 minutes, achieved complete extraction across all abstracts, and produced concise, synthesized outputs that are well suited for downstream analysis. Llama also performed well and represents a viable alternative, particularly for generating concise outputs; however, its longer processing time (approximately 40-minute for the full corpus) may limit scalability for larger-scale applications. In contrast, Athena consistently exhibited a tendency to reproduce source text directly rather than synthesize information into concise, field-specific summaries. Although verbatim extraction may preserve surface-level textual fidelity, it reduces the practical utility of the generated outputs by limiting information distillation, occasionally incorporating information from unrelated characteristics into individual fields, and increasing output length without providing additional analytical value. Collectively, these findings suggest Gemma and Llama are better suited for this use case, where synthesized, concise, and structured outputs are needed to support large-scale literature review and downstream analysis.

Human evaluation of 50 randomly selected abstracts supported the overall model rankings and highlighted the limitations of relying solely on automated metrics to assess extraction quality. Although Token F1 scores ranked Llama higher than Gemma, expert evaluation favored Gemma by a substantial margin, with 68.0% of Gemma-generated outputs rated as good compared with 36.0% of Llama’s outputs. This discrepancy suggests that token-level agreement with a majority-vote reference does not fully capture the dimensions of quality most relevant to end users, including semantic correctness, appropriate field scoping, concise synthesis, and avoidance of hallucination. These findings underscore the importance of incorporating expert evaluation alongside automated metrics when assessing LLM performance for specialized biomedical information extraction tasks. Such evaluation is particularly important in biomedical application, where extraction errors, such as misattributed clinical outcomes or unsupported study characteristics, could affect downstream evidence synthesis, knowledge representations, or research conclusions.

Differences in extraction completeness across characteristics likely reveal both variation in abstract reporting practices and differences in model interpretation. Characteristics such as disease name, study purpose, study type, and clinical outcomes were extracted from nearly all abstracts by all models, suggesting that these elements are consistently reported in NHS literature and are relatively straightforward for LLMs to identify. In contrast, characteristics with higher missing output rates, such as exclusion criteria, which were absent in 40.3% to 95.2% of abstracts across models, likely reflect information that is less commonly reported in abstracts and may instead appear only in full-text publications. Fields with greater variation in missing output across models, such as treatments received (3.6% to 50.1%) and data collection period (5.2% to 54.8%), may indicate both inconsistent reporting practices in NHS abstracts and differences in how models interpret and extract these characteristics. Similarly, mapping extracted terms to controlled vocabularies revealed comparable trends across models: GARD and RxNorm mapping rates were relatively high and consistent across models, whereas HPO mapping rates were substantially lower. This finding suggests that phenotype extractions may require additional normalization strategies to improve alignment between free-text clinical descriptions and standardized ontologies. Overall, our findings demonstrate that open-source LLMs can be effectively applied to specialized biomedical information extraction tasks. As LLM capabilities continue to improve, these approaches have the potential to accelerate the extraction and structuring of information from biomedical literature, improve the accessibility to natural history study evidence, and support large-scale rare disease research.

Despite these promising results, we acknowledge several limitations. First, as proof of concept, our evaluation was limited to PubMed abstracts rather than full-text articles. As a result, extraction completeness may have been constrained for study characteristics that are often underreported in abstracts, such as exclusion criteria, data collection period, and detailed treatment information. In addition, we did not analyze natural history study-related clinical trial records, which may contain more practical and structured information about study design, eligibility criteria, enrollment status, outcome measures, and data collection plans. Future work incorporating full-text articles and clinical trial records may improve extraction completeness and provide a more comprehensive assessment of natural history study characteristics. Second, our framework primarily focused on qualitative study descriptors and expert review of the extracted information, rather than systematic extraction of quantitative results, such as, p-values, hazard ratios, and other statistical measures. Extending the framework to capture numerical outcomes would further enhance its utility but would require additional prompt engineering, validation, and quality control procedures to ensure statistical values are accurately extracted and correctly attributed to their corresponding study context. Finally, the models evaluated through the NCATS computational infrastructure differed in parameter count: Llama (70 billion parameters), Gemma (27 billion parameters), and Athena (13 billion parameters). Differences in model scale may influence reasoning capabilities, knowledge representation, output quality, runtime, and computational requirements. Future evaluations should consider comparisons among models with similar parameter sizes or develop efficiency-normalized evaluation metrics that jointly account for extraction quality, processing time, and computational cost across model scales.

### Conclusions

This study demonstrates the feasibility of using open-source LLMs for automated information extraction from NHS abstracts. Through an iterative prompt engineering strategy and comparative evaluation of three locally available models, it showed that LLMs can efficiently extract structured characteristics of rare disease studies at scale with high completion rates. Among the evaluated models, Gemma demonstrated the best overall balance of runtime efficiency, extraction completeness, and expert-rated output quality, while Llama also performed well but required longer processing time. These findings support the potential of locally deployed open-source LLMs to improve access to NHS evidence, reduce the burden of manual curation, and accelerate rare disease research. Future efforts incorporating full-text articles, clinical trial records, quantitative extraction of outcomes, and broader benchmarking across additional models will further strengthen this framework and expand its applicability for large-scale biomedical literature analysis.

## List of abbreviations

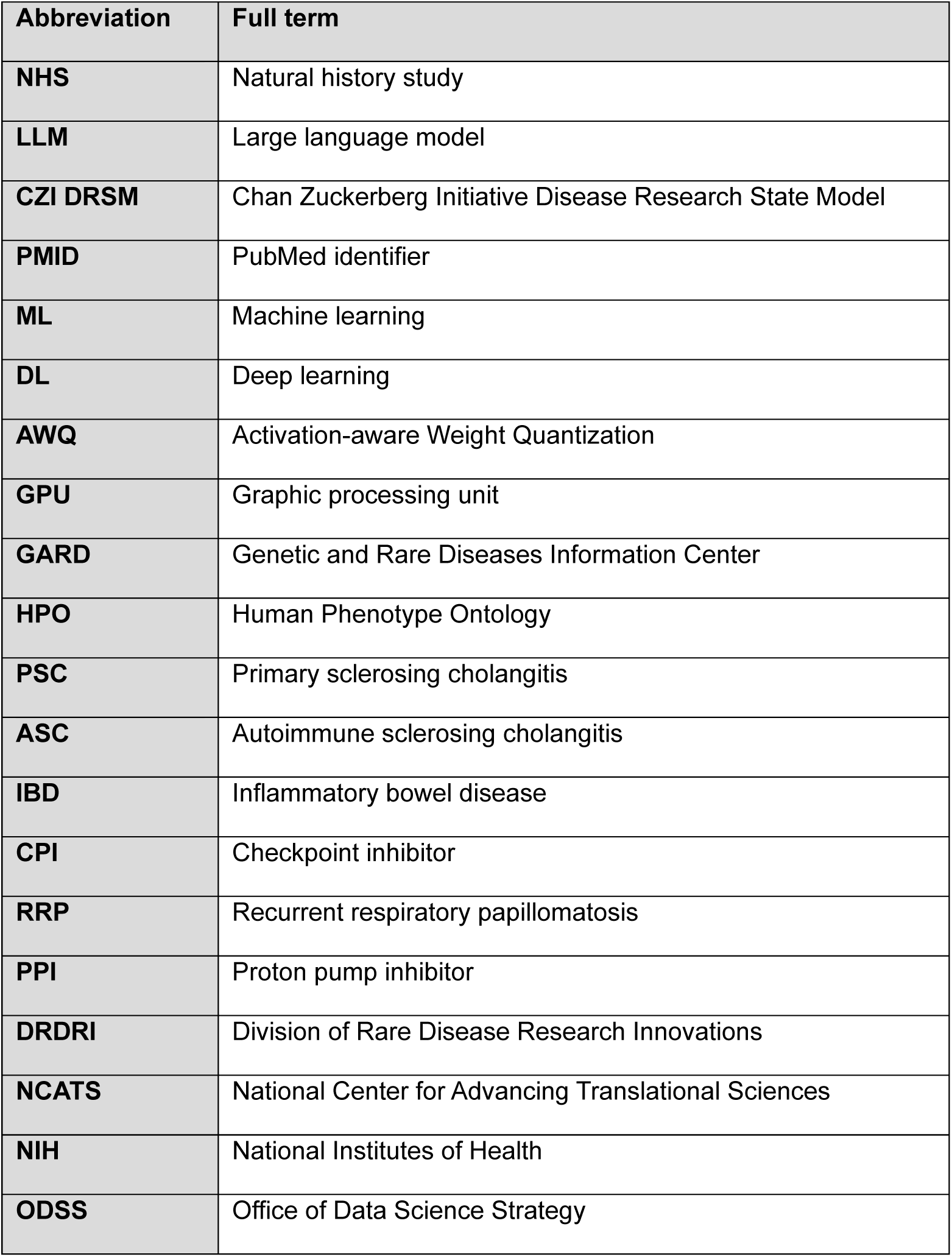

## Declarations

### Ethics approval and consent to participate

Not applicable.

### Consent for publication

Not applicable.

### Code and Data Availability

The information extraction pipeline and analysis code used in this study are available on GitHub.[29] The PubMed abstract corpus used in this study is the publicly available Chan Zuckerberg Initiative Disease Research State Model (CZI DRSM) corpus.[18]

### Contributions

KL: performed the study and wrote the manuscript; ES: as a rare disease expert, guided NHS-focused research questions and manually compared LLM performance; QZ: conceived and supervised this study and wrote the manuscript. All authors reviewed and approved the manuscript.

## Data Availability

All data and code produced are available online at GitHub in https://github.com/ncats/RDAS/tree/nhs_extraction_dockerization/NHS_info_extraction.

## References

1. Orphan Drug Act of 1983, Pub. L. No. 97-414, 21 U.S.C. § 360bb.

2. Haendel M, Vasilevsky N, Unni D, et al. How many rare diseases are there? Nat Rev Drug Discov. 2020;19(2):77–78. doi:10.1038/d41573-019-00180-y

3. Nguengang Wakap S, Lambert DM, Olry A, et al. Estimating cumulative point prevalence of rare diseases: analysis of the Orphanet database. Eur J Hum Genet. 2020;28(2):165–173. doi:10.1038/s41431-019-0508-0

4. National Human Genome Research Institute. Rare Diseases FAQ. https://www.genome.gov/FAQ/Rare-Diseases

5. GARD - Genetic and Rare Diseases Information Center. About GARD. National Center for Advancing Translational Sciences, NIH. https://rarediseases.info.nih.gov/about

6. Boulanger V, Schlemmer M, Rossov S, Seebald A, Gavin P. Establishing Patient Registries for Rare Diseases: Rationale and Challenges. Pharm Med. 2020;34(3):185–190.

7. U.S. Food and Drug Administration. Rare Diseases: Natural History Studies for Drug Development. Draft Guidance for Industry. Docket No. FDA-2019-D-0481; March 2019.

8. Augustine EF, Adams HR, Mink JW. Clinical trials in rare disease: challenges and opportunities. J Child Neurol. 2013;28(9):1142–1150. doi:10.1177/0883073813495959

9. European Medicines Agency. Guideline on Registry-Based Studies. EMA/502388/2020; October 2021.

10. Liu J, Barrett JS, Leonardi ET, Lee L, Roychoudhury S, Chen Y, Trifillis P. Natural History and Real-World Data in Rare Diseases: Applications, Limitations, and Future Perspectives. J Clin Pharmacol. 2022;62(Suppl 2):S38–S55. doi:10.1002/jcph.2134

11. Aronson AR. Effective mapping of biomedical text to the UMLS Metathesaurus: the MetaMap program. Proc AMIA Symp. 2001:17–21.

12. Tang B, Cao H, Wu Y, Jiang M, Xu H. Recognizing clinical entities in hospital discharge summaries using Structural Support Vector Machines with word representation features. BMC Med Inform Decis Mak. 2013;13(Suppl 1):S1. doi:10.1186/1472-6947-13-S1-S1

13. Bundschus M, Dejori M, Stetter M, Tresp V, Kriegel HP. Extraction of semantic biomedical relations from text using conditional random fields. BMC Bioinformatics. 2008;9:207. doi:10.1186/1471-2105-9-207

14. Lee J, Yoon W, Kim S, et al. BioBERT: a pre-trained biomedical language representation model for biomedical text mining. Bioinformatics. 2020;36(4):1234–1240. doi:10.1093/bioinformatics/btz682

15. Gu Y, Tinn R, Cheng H, et al. Domain-specific language model pretraining for biomedical natural language processing. ACM Trans Comput Healthc. 2021;3(1):1–23. doi:10.1145/3458754

16. Brown TB, Mann B, Ryder N, et al. Language models are few-shot learners. Adv Neural Inf Process Syst. 2020;33:1877–1901.

17. Agrawal M, Hegselmann S, Lang H, Kim Y, Sontag D. Large language models are few-shot clinical information extractors. Proceedings of the 2022 Conference on Empirical Methods in Natural Language Processing (EMNLP). 2022:1998–2022.

18. Chen K, Ao M, Moon S, Burns G, Zhu Q. Machine Learning-Based Identification of Natural History Studies in Rare Diseases: A Step toward Understanding Disease Development and Outcome. J Rare Dis (Berlin). 2025;4(1). doi:10.1007/s44162-025-00115-9

19. TheBloke. Athena V3 - AWQ [Internet]. Hugging Face; [cited 2026 Aug 5]. Available from: https://huggingface.co/TheBloke/Athena-v3-AWQ. Based on Athena V3 by IkariDev and Undi95, available from: https://huggingface.co/IkariDev/Athena-v3.

20. Lin J, Tang J, Tang H, Yang S, Chen WM, Wang WC, Xiao G, Dang X, Gan C, Han S. AWQ: Activation-aware Weight Quantization for LLM Compression and Acceleration. Proceedings of Machine Learning and Systems (MLSys). 2024;6. arXiv:2306.00978.

21. Kamath A, et al. (Gemma Team, Google DeepMind). Gemma 3 Technical Report. arXiv:2503.19786. Published March 12, 2025.

22. Grattafiori A, et al. (Llama Team, Meta AI). The Llama 3 Herd of Models. arXiv:2407.21783. Published July 31, 2024.

23. Valentino PL, Wiggins S, Harney S, Raza R, Lee CK, Jonas MM. The Natural History of Primary Sclerosing Cholangitis in Children: A Large Single-Center Longitudinal Cohort Study. J Pediatr Gastroenterol Nutr. 2016;63(6):603–609. doi:10.1097/MPG.0000000000001368

24. Kwon W, Li Z, Zhuang S, Sheng Y, Zheng L, Yu CH, Gonzalez JE, Zhang H, Stoica I. Efficient Memory Management for Large Language Model Serving with PagedAttention. Proceedings of the 29th ACM Symposium on Operating Systems Principles (SOSP ‘23). 2023:611–626. arXiv:2309.06180.

25. Köhler S, Gargano M, Matentzoglu N, et al. The Human Phenotype Ontology in 2021. Nucleic Acids Res. 2021;49(D1):D1207–D1217. doi:10.1093/nar/gkaa1043

26. Nelson SJ, Zeng K, Kilbourne J, Powell T, Moore R. Normalized names for clinical drugs: RxNorm at 6 years. J Am Med Inform Assoc. 2011;18(4):441–448. doi:10.1136/amiajnl-2011-000116

27. Gomes F, Lorigan P, Woolley S, Foden P, Burns K, Yorke J, Blackhall F. A prospective cohort study on the safety of checkpoint inhibitors in older cancer patients – the ELDERS study. ESMO Open. 2021;6(1):100042. doi:10.1016/j.esmoop.2020.100042

28. Benedict PA, Ruiz R, Yoo M, Verma A, Ahmed OH, Wang B, Dion GR, Voigt A, Merati A, Rosen CA, Amin MR, Branski RC. Laryngeal distribution of recurrent respiratory papillomatosis in a previously untreated cohort. Laryngoscope. 2018;128(1):138–143. doi:10.1002/lary.26742

29. National Center for Advancing Translational Sciences. NHS Information Extraction Pipeline [Internet]. GitHub. Available from: https://github.com/ncats/RDAS/tree/nhs_extraction_dockerization/NHS_info_extraction.

